# Study protocol for a cluster randomized controlled trial for 16 weeks to assess the effect of almond consumption on glycemic control and gut health in prediabetes adults in rural settings of India

**DOI:** 10.1101/2023.06.21.23291579

**Authors:** Ruchi Vaidya, Nayanjeet Chaudhury, Pramila Kalra, Sreekumaran Nair, Bellur S. Prabhakar

## Abstract

**Introduction:** Almonds have prebiotic potential to maintain gut health and regulate glycemia. Western studies have shown their positive effects in preventing non-communicable diseases like diabetes and CVDs. However, there is a lack of well-designed studies involving Asian Indians, who have a higher predisposition to diabetes due to their unique ‘Asian phenotype’. Therefore, this study aims to evaluate the impact of almond supplementation on glycemic control and gut health in prediabetic adults in rural India through a randomized clinical trial.

**Methods and Analysis:** A parallel cluster randomized controlled trial with 178 prediabetic participants aged 20-50, both genders, with a BMI of 18.9-25 kg/m2, will be conducted in rural areas of Chikkaballapur, Kolar, and rural Bangalore districts in India. The intervention group will receive 56g of almonds as mid-morning snacks for 16 weeks, while the control group will receive cereal pulse based traditional isocaloric snacks under the closed supervision of the study investigators. Anthropometry, clinical, and biochemical parameters will be measured at 0, 8th, and 16th weeks, and a subgroup of 120 participants will undergo gut health analysis. GLP 1 analysis will be conducted on 30 participants at 0 and 16th week. Statistical analysis will be performed using SPSS for Windows V 27.0, and both ITT and per-protocol analyses will be conducted.

**Ethics and Dissemination:** Ethics approval was obtained from the Institutional Ethics Committee at Ramaiah Medical College, Bangalore, Karnataka, India (DRPEFP7672021). Results from this trial will be disseminated through publication in peer-reviewed journals, national and international presentations.

**Trial Registration Number:** The trial is registered in the clinical trial registry of India (CTRI/2023/03/050421).

**ARTICLE SUMMARY:** Strengths and Limitations:

- The study is the first field-based trial in rural settings to assess the effect of almond consumption versus traditional cereal pulse-based snacks on prediabetes adults’ glycemic control and gut health.
- The compliance management is crucially designed by performing an intervention on one-to-one closed supervision, assessment of serum tocopherol levels, and regular follow up of any adverse events.
- The upcoming metagenomic analysis and the gut metabolites like short chain fatty acids (SCFA) and GLP 1 will significantly advance our understanding of the impact of almonds on the gut health of prediabetes adults.
- A potential limitation of the study is that it will not be feasible to follow the participants after the post intervention period.

## INTRODUCTION

India is one of the epicentres of diabetes, with 77 million people living with diabetes and 33 million with prediabetes [1]. The so-called “South Asian” or “Asian-Indian” phenotype makes this ethnic group more susceptible to type 2 diabetes and premature coronary artery disease than white Caucasians [2]. Despite the lower prevalence of generalized obesity, the phenotype is characterized by increased insulin resistance, higher diabetes rates, central adiposity, and dyslipidaemia, with raised serum triglycerides and low levels of high-density lipoprotein (HDL) cholesterol [3]. While genetic factors might contribute a little to the South Asian phenotype, the current diabetes epidemic is fuelled predominantly by lifestyle, mainly unhealthy diet and physical inactivity [4].

Recent evidence suggests a link between human metabolic health and bacterial populations in the gut, and it is primarily controlled by the nutritional quality of the diet. Colonic microbiota can be modulated positively or negatively by different lifestyle and dietary factors and impact the risk of developing obesity and lifestyle related non-communicable diseases (NCDs) (e.g., diabetes, CVD, and metabolic syndrome features) and infectious diseases. Metabolic condition like diabetes alters the gut microbial composition [5]. Developing strategies to modify the gut microbiota to control glycemic and lipemic regulations is especially important.

Microbial products, such as SCFAs (Short chain fatty acids), may affect metabolism by regulating appetite, lipogenesis, gluconeogenesis, inflammation, and producing Glucagon like peptide 1 (GLP-1) to regulate glucose metabolism. Whereas intestinal dysbiosis can change the functioning of the intestinal barrier and the gut-associated lymphoid tissues (GALT), which activate inflammatory pathways and may contribute to the development of insulin resistance, hyperglycemia and cardiometabolic risks [6].

Almonds are commonly consumed nuts in India in either raw or roasted form. There is a growing body of randomized controlled trials (RCTs) that is emerging to support almond’s role in promoting healthy microbiota [7-11]. Almonds contain considerable amounts of potential prebiotic components. The pectic substances encasing the cellulose microfibrils are a significant component of almond skin cell walls, with small hemicellulose such as xyloglucan and alpha glucan. A study on healthy Chinese adults indicated that almond supplementation improved intestinal microbiota profile (significant increase in *bifidobacteria spp* and *lactobacilli spp*) and modified intestinal bacterial activities (significant increase in fecal β-galactosidase activity and decreases in fecal β-glucuronidase, nitroreductase and azoreductase activities) [7]. Along with the prebiotic quality, almonds are rich in healthy unsaturated fats, fibre, proteins, vitamin E and B-vitamins, calcium, magnesium and copper, and phytosterols and polyphenols, and low in available carbohydrates, which can support various healthy biomarkers associated with the prevention and management of chronic diseases [11]. The prevalence of prediabetes among adults over 20 years in Karnataka is 11.7% (10.6% rural and 14.2% urban) [12]. However, the spread of this epidemic to economically disadvantaged sections of society is a matter of great concern in India, where most expenses for the treatment of diabetes are borne out of pocket by patients.

Given that 65% of India’s population resides in rural areas, even a minor increase in the rural prevalence of diabetes will translate into several millions of individuals requiring chronic care. Considering the lack of awareness and poor health care services in rural settings, it is inevitable to strategize preventive measures to halt the rise in diabetes and related complications. Therefore, there is an unmet need to evolve dietary strategies from well-conducted randomized clinical trials to mitigate the health and economic burden of diabetes. This article explains the study design and methods of a randomized trial to evaluate the effect of 56 g almond supplementation for 16 weeks among rural Indian prediabetes adults with the following objectives:

## STUDY OBJECTIVES

### Primary objective

The primary objective of the study is to evaluate the effect of almond consumption for 16 weeks on glycemic control (HbA1c levels) among rural Indian adults with prediabetes compared to that of consumption of traditional cereal-pulse based snack. We hypothesize that consuming 56g of almonds per day for 16 weeks may induce colonization of good bacteria and short chain fatty acid production (SCFA) in the gut, which plays a vital role in the management of blood glucose levels in prediabetes adults compared to the usual cereal pulse-based snack consumption.

### Secondary objectives

The secondary aim is to determine the effect of almond consumption on the colonization of beneficial gut microflora and fecal short chain fatty acid (SCFA). We also aim to understand the association between gut bacterial activities and blood glucose levels in rural Indian adults with prediabetes.

### Tertiary objectives

The tertiary objective is to evaluate whether almond consumption over 16 weeks compared to traditional mid-morning snacks results in any beneficial changes in anthropometric measurements, in particular, waist circumference and body weight and whether it improves liver enzyme profiles, improves inflammatory biomarkers, lipid profile, fasting insulin, HOMA IR, serum creatinine and gut hormones like GLP-1. The study also aims to observe any beneficial association in their physical activity pattern and nutrient intake upon consumption of almonds over traditional cereal pulse-based snacks for a period of 16 weeks.

## METHODS AND ANALYSIS

### Study design

The study is designed as a 16-week cluster randomised parallel-arm controlled dietary intervention in the rural settings of India. It is a superior trial framework with an equal allocation ratio of the participants in both arms. The trial will be conducted in the research facilities of the Ramaiah International Centre for Public Health Innovations, Bangalore, Karnataka, India. The Standard Protocol Items: Recommendations for Interventional Trials guidelines (SPIRIT) were used in the development of this protocol [13].

### Study settings

It is a community-based trial that will be carried out in a free living population residing in rural settings of Karnataka (South India). Almost 65% of Indians reside in rural settings, and considering the lack of awareness and poor health care services, the study is designed to treat rural adults and provide them with an accessible and culturally appropriate dietary approach for the prevention of diabetes. RICPHI manages a multi-district community outreach program, with subdistricts as clusters for intervention on chronic diseases. Therefore, the given population shall be recruited from the villages of *Chikkaballapur, Kolar* and *Rural Bangalore* districts of Karnataka, which are within the same geography where RICPHI’s community outreach program exists.

### Participant eligibility

The study includes participants in the age group 20-50 years with a BMI range 18.9-25 kg/m2. As the chosen population is known to manifest the Asian Indian phenotype, the prevalence of diabetes is high among adults more than 20 years in both normal and overweight/obese individuals [3]. Moreover, this age range ensures that physical maturity has been achieved. Also, the prevalence of chronic health conditions that fit our exclusion criteria for volunteer participation in the study is low in this age range. Therefore, the study participants should be willing to be a part of the study, with no nut allergies, fasting plasma glucose levels between 100-125 mg/dl and/or random blood glucose levels between140 -199 mg/dl, HbA1c levels between 5.7 to 6.4%, no comorbidities such as thyroid, diabetes, chronic kidney disease, alcoholic liver disease, or any known case of cardiovascular disease, or any chronic illness, not taking medications (steroids and beta-blockers, antibiotics) known to influence metabolism and appetite and should not be a chronic alcohol dependent. All eligible participants will be screened and enrolled after obtaining informed consent.

Recruitment/Screening: The study plans to select villages for the study based on their population size and proximity to the head office. The team will meet District Health Officers, Medical officers and primary health care workers in the chosen areas to discuss the program and secure their consent. To ensure good participation, the team will first perform sensitization and rapport building exercises to develop trust with the community. The study will invite adults aged between 20 and 50 years who have lived in the selected village for at least six months to participate in the Level 1 screening to identify their risk of prediabetes. To understand the risk of prediabetes in this population, the study will use the Indian Diabetes Risk Score (IDRS)[14] and Prediabetes Risk Score (PRESS) [15]. Those with high and moderate risk group will undergo HbA1c analysis in Level 2 screening to diagnose prediabetes. The study will conduct further investigations on confirmed prediabetes adults to check for other comorbidities like liver disorders, kidney disorders, thyroid, cardiac health, inflammatory markers and any other comorbidities. After obtaining informed consent, those who meet all the inclusion criteria will be enrolled in the study.

### Sample size calculation

Considering 0.65 as the minimal clinical significant difference in mean HbA1c levels, a standard deviation of 1, for 80% power and 5% level of significance, the minimum number of participants required for the study after considering a 20% dropout rate has been estimated to be 148. Considering a design effect of 1.2, the minimum number of total participants required is 178. The study will identify 20 clusters, with 10 for the control and intervention groups. Each cluster will define one anganwaadi worker’s (AWW) area as a unit. An AWW in an Indian rural setting covers 1000 population. The cluster size will be limited to 10-12 participants.

### Randomization, Allocation concealment and sequence generation

Randomization will be performed by an independent biostatistician using a computer-generated random number sequence that will be sealed in numbered opaque envelopes [16]. A staff member independent of the study outcome assessments and statistical analysis will perform the treatment allocation and keep the sealed envelopes in a secure location with access limited to authorised personnel. As the participants are consuming whole foods, which are easily identified, the participants and staff involved in the intervention cannot be blinded.

### Pre-Intervention

#### Snack consumption pattern

To understand the snack consumption pattern of the community in the study sites, a small study was conducted using focus group discussions and individual interviews. The results showed that most participants consumed snacks around mid-morning and evening, and they usually consumed tea or coffee thrice or four times a day with cereal pulse-based snacks, either homemade or purchased from local shops or bakeries.

#### Acceptability of almonds

To gauge the acceptability of almonds as a snack, ten adults were given 56 grams of almonds for three consecutive days, and their preference for consuming almonds was analysed. They all enjoyed the taste of almonds and preferred to eat them as a part of mid-morning snacks. In a focus group discussion, participants suggested gathering in one place to eat the almonds together instead of taking the packets home.

#### Isocaloric control food selection

The availability of snacks in the local grocery shops of the study areas was also studied. The most popular snacks were analysed for their macronutrient content and ingredients. Snacks that did not contain nuts were further analysed for their macronutrient content. Three popularly consumed ready-to-eat snacks were selected as control food. The comparison of their nutrient composition is shown in Table 1.

**Table 1:** - Nutrient content of Intervention food & Control food selection.

| Macro-Nutrients | California Almonds | Control Food 1 | Control Food 2 | Control Food 3 |
| --- | --- | --- | --- | --- |
| *Moisture %* | 4.3 | 1.4 | 3.3 | 3.3 |
| *Protein (g %)* | 30.7 | 7 | 3 | 17.6 |
| *Total Fat (g%)* | 48.3 | 39.4 | 33.6 | 18.8 |
| *Total Carbohydrates (g%)* | 6.4 | 50.4 | 54.9 | 55.3 |
| *Crude Fibre (g%)* | 8.5 | 0.5 | 2.3 | 2 |
| *Total Ash (%w/w)* | 1.8 | 1.3 | 2.9 | 3 |
| *Total Calories (%kcal)* | 583.1 | 584.2 | 534 | 405.8 |
| *Isocaloric amount of the control food* | 56g | 56g | 61g | 80g |

### Intervention

The study intervention plan is summarized in Table 2. The eligible participants will be enrolled in the study after obtaining informed consent. Prior to the study intervention, we will perform a run-in period of 2 weeks. The intervention arm participants will receive California almonds as mid-morning snacks. In contrast, the control group will receive cereal pulse-based traditional snacks with calories equivalent to the almonds consumed by the intervention group daily for 16 weeks. The almonds to be supplemented will be California almonds with un-blanched kernels and brown skin intact, and the participants’ should consume 56g of almonds daily for 16 weeks as a replacement for their traditional cereal pulse-based mid-morning snack. Field workers will closely supervise their intake. The participants will replace approximately 340 calories of their regular diet with 56g of California almonds. Throughout the study period, participants of both groups will be instructed to adhere to their regular dietary and physical activity practices, and we will monitor this every week using a mobile app-based monitoring system.

**Table 2:**
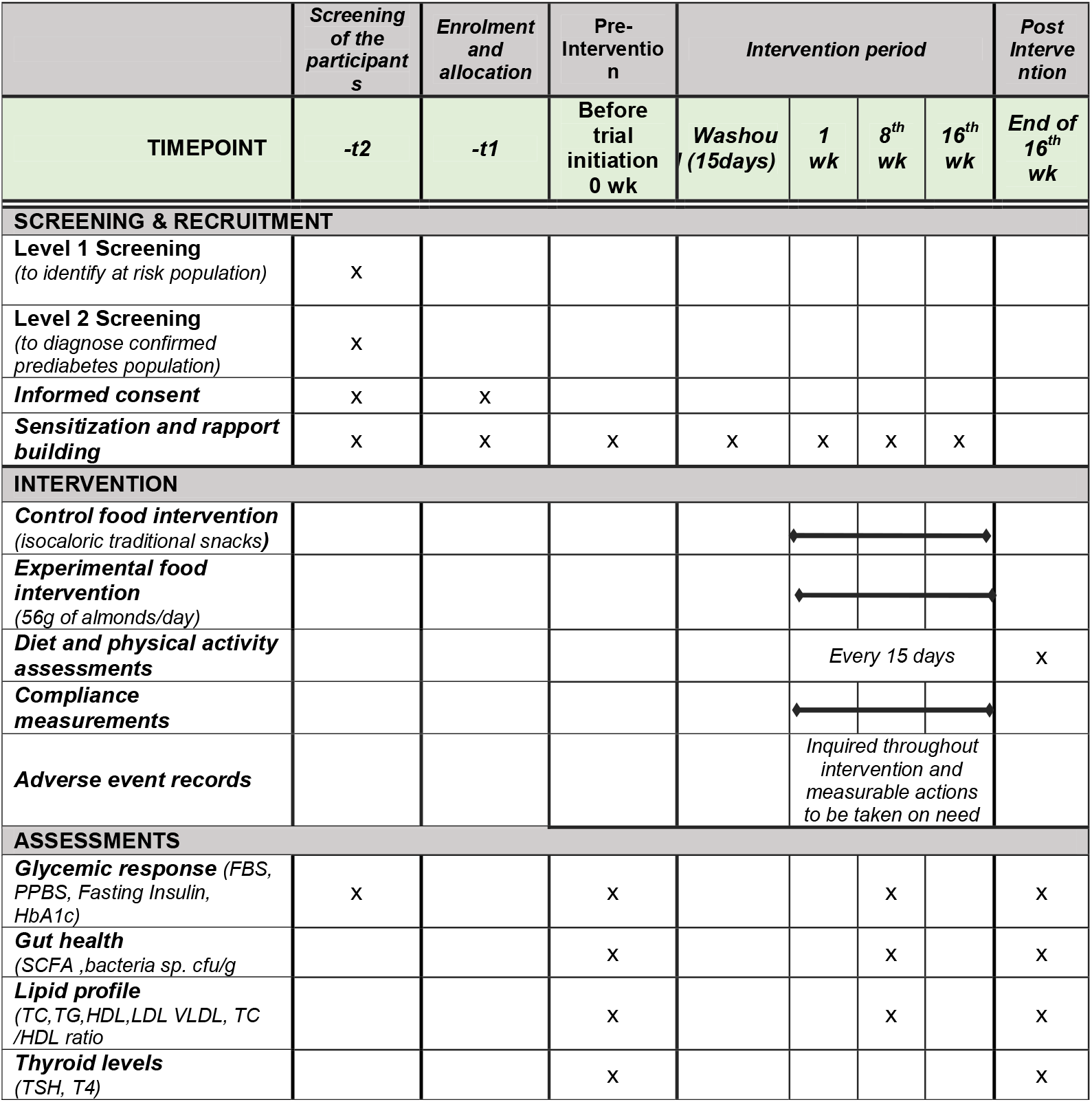
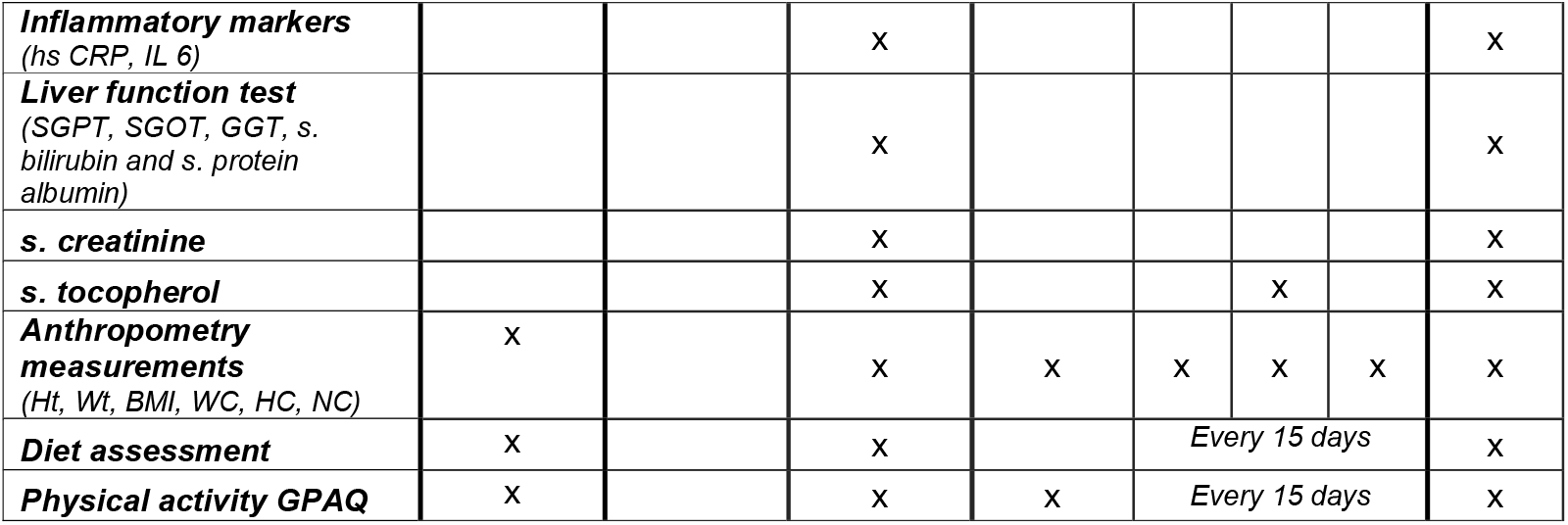
Study protocol, Intervention and assessments plan for the trial.

|  | Screening<br>of the<br>participant<br>s | Enrolment<br>and<br>allocation | Pre-Interventio<br>n | Intervention period |  |  |  | Post<br>Interven<br>tion |
| --- | --- | --- | --- | --- | --- | --- | --- | --- |
| TIMEPOINT | -t2 | -t1 | Before<br>trial<br>initiation<br>0 wk | Washou<br>t<br>(15days) | 1<br>wk | 8<sup>th</sup><br>wk | 16<sup>th</sup><br>wk | End of<br>16<sup>th</sup><br>wk |
| SCREENING & RECRUITMENT |  |  |  |  |  |  |  |  |
| Level 1 Screening<br>(to identify at risk population) | X |  |  |  |  |  |  |  |
| Level 2 Screening<br>(to diagnose confirmed<br>prediabetes population) | X |  |  |  |  |  |  |  |
| Informed consent | X | X |  |  |  |  |  |  |
| Sensitization and rapport<br>building | X | X | X | X | X | X | X |  |
| INTERVENTION |  |  |  |  |  |  |  |  |
| Control food intervention<br>(isocaloric traditional snacks) |  |  |  |  |  |  |  |  |
| Experimental food<br>intervention<br>(56g of almonds/day) |  |  |  |  |  |  |  |  |
| Diet and physical activity<br>assessments |  |  |  |  | Every 15 days |  |  | X |
| Compliance<br>measurements |  |  |  |  |  |  |  |  |
| Adverse event records |  |  |  |  | Inquired throughout<br>intervention and<br>measurable actions<br>to be taken on need |  |  |  |
| ASSESSMENTS |  |  |  |  |  |  |  |  |
| Glycemic response (FBS,<br>PPBS, Fasting Insulin,<br>HbA1c) | X |  | X |  |  | X |  | X |
| Gut health<br>(SCFA ,bacteria sp. cfu/g) |  |  | X |  |  | X |  | X |
| Lipid profile<br>(TC,TG,HDL,LDL VLDL, TC<br>/HDL ratio |  |  | X |  |  | X |  | X |
| Thyroid levels<br>(TSH, T4) |  |  | X |  |  |  |  | X |
| **Inflammatory markers**<br>(hs CRP, IL 6) |  |  | X |  |  |  |  | X |
| **Liver function test**<br>(SGPT, SGOT, GGT, s. bilirubin and s. protein albumin) |  |  | X |  |  |  |  | X |
| **s. creatinine** |  |  | X |  |  |  |  | X |
| **s. tocopherol** |  |  | X |  |  | X |  | X |
| **Anthropometry measurements**<br>(Ht, Wt, BMI, WC, HC, NC) | X |  | X | X | X | X | X | X |
| **Diet assessment** | X |  | X |  | Every 15 days |  |  | X |
| **Physical activity GPAQ** | X |  | X | X | Every 15 days |  |  | X |

### Investigations

#### Glycemic response

Blood will be drawn at 0, 8, and 16 weeks to measure fasting blood glucose levels, HbA1c, and insulin levels to analyse the glycemic response.

#### Gut health

The fecal samples will be collected in a sub group of 120 participants at baseline (0), midline (8 weeks) and end line (16 weeks) to measure the short chain fatty acid levels and beneficial gut microbiota counts. Fecal samples will be assessed for short chain fatty acid composition (GCMS method) and gut microbiota sequencing (PCR) at baseline, midline and at the end of the study period. The blood and fecal samples will be collected and stored in sterilized temperature-controlled storage boxes and transported to the laboratories for testing within 6-8 hours.

#### Other tests

The metabolic markers such as lipid profile, liver function tests (SGPT, SGOT, GGT, s. bilirubin and s. protein albumin), s creatinine and s, urea, thyroid stimulating hormone (TSH) anf T4 harmone levels and inflammatory markers (hs CRPP and IL-6) will be measured at baseline and endline.

In a sub-group of 60 participants, active plasma glucagon like peptide 1 (GLP-1) levels will be estimated immediately after the consumption of midmorning snacks at baseline and end line of the study. For this, blood samples will be drawn at the following periods after almond consumption: 0 (Immediately after consumption and at 10 and 30 minutes.

### Anthropometry

Their anthropometric measurements (body weight, body height, BMI, waist circumference, neck circumference and hip circumference) and systolic and diastolic blood pressure will be measured at the baseline (0), midline (8 weeks) and the end of the study (16 weeks).

### Dietary and physical assessment

The dietary intake and physical activity pattern will be monitored periodically using validated tools, 24-hour dietary recall and the Global Physical activity questionnaire (GPAQ). The mobile phone based electronic data capture tool will be specially developed based on the tools.

### Adverse events

Adverse events will be recorded in the case report form and will be reported to the Institutional Ethics Committee. Side effects will be unlikely, but if any severe side effects due to the intervention are detected, participants will be withdrawn from the study. Adverse events that lead to withdrawals will be reported in future publications. We do not intend to analyse adverse events formally.

In order to ensure a high degree of compliance, incentives and personal health & fitness services will be provided to the study participants in both groups at the end of the study for a month by a team of certified fitness trainers, nutritionists and physicians via an official agreement with the community heads of those rural settings.

### Data management

An electronic Management Information System (MIS) application has been developed to monitor the field activities, enable real time data collection and support the trial implementation team with dashboards to assist real time decisions. The data will be collected in the field in a physical form and electronic form into a central database. The platform has an inherent capability to create a unique ID for each participant’s data being submitted, which will be used to identify their electronic and paper-based data and biological samples. A password-protected computer database, accessible by the researchers only, will store participant identifiers (e.g., name, email address, phone number) and their associated code. Paper-based data will be stored securely at the Ramaiah Institute of Public Health Innovations for five years, after which it may be destroyed. Biological samples will be stored at −80°C freezer; Samples will be stored for up to 3 years from the collection date and disposed of accordingly after that time.

### Statistical analysis plan

Statistical analysis will be performed using SPSS for Windows V 27.0. Intention-to-treat (ITT) and per-protocol analyses (for those who achieve a minimum of 80% compliance with test food consumption) will be performed. Bonferroni post hoc tests will be performed where main effects are identified to identify significant differences between means (p <0.05). While the ITT analysis will be the primary analysis, the per-protocol analysis will allow us to decipher whether the effects are due to participants being compliant with consuming test foods. We will also run sub-analyses. Descriptive statistics (means and standard deviations, quartiles for continuous variables and frequencies and percentages for categorical variables) will be reported. Bivariate comparisons, using chi-square tests for categorical, one-way ANOVA/ Kruskal Wallis test, independent student t test/ Mann Whitney U test for continuous variables based on the normality of the data, will be conducted. The impact of the intervention will be assessed using repeated measures ANOVA, Analysis of variance with Tukey’s posthoc test will be used for intra-and inter-group comparisons. A two-tailed p-value < 0.05 will be considered statistically significant.

A data safety monitoring board (DSMB) is formed to monitor participant safety, data quality and progress of the ABC Trial program. In addition, the DSMB will review study data periodically to evaluate the study’s safety, conduct, scientific validity, and data integrity. The members of DSMB are selected by the Advisory panel of the project.

### Ethics and Dissemination

Ethics approval was obtained from the Institutional Ethics Committee at Ramaiah Medical College, Bangalore, Karnataka, India (DRPEFP7672021). Participants will receive a copy of their results (with the exception of their faecal sample testing as well as a summary of the study findings. Results from this trial will be disseminated through publication in peer-reviewed journals, national and international presentations. The trial is registered in the clinical trial registry of India (CTRI/2023/03/050421).

## DISCUSSION

Randomized controlled trials (RCTs) have demonstrated the effectiveness and cost-efficiency of medical nutrition therapy in improving the metabolic outcomes of patients, as evidenced by numerous studies [17]. It is increasingly recognized that nutrition science would benefit from more robust and well-conducted RCTs to evaluate the impact of nutrients and foods on clinical outcomes, which can complement findings from prospective cohorts and metabolic studies. Each of these research approaches has its advantages and limitations, and consistent findings can significantly contribute to developing dietary guidelines and their successful implementation. In India, where cardiovascular disease and diabetes are known to increase disability-adjusted life years,[18] it is crucial to establish nutrition interventions that can help prevent these diseases, particularly through their impact on modifiable risk factors such as hyperglycemia. The consumption of nuts aligns with this urgent need.

Almonds have beneficial effects on various health markers, including a lipid profile, atherogenic indices, endothelial function, inflammatory markers, glycemic control, HbA1c, and blood pressure, as supported by research [19,20]. However, there is a lack of studies on the effects of almond consumption on glucose responses and gut health in India. A short-term intervention trial lasting up to 4 weeks has shown the beneficial effect of almond consumption on glycemic response and cardio-metabolic risks in healthy and at-risk adolescents and young adults in India [21].

A long-term pre-post clinical trial study was carried out among 50 North Indian subjects with Type 2 diabetes from New Delhi, India. The middle-aged adults (mean age 46 years, mean BMI 29) were enrolled for a 3-week run-in diet, followed by a post-run-in diet which included 20% energy from raw almonds (about 60 g/day) for 24 weeks. Compared to the run-in diet, the almond-enriched diet significantly improved TC, LDL-C, VLDL-C, triglycerides, HbA1c, and hs-CRP, enhancing pulse wave velocity [22,23]. However, the study has not explored the mechanisms and prebiotic potential of almonds and their role in the management of glycemic and lipemic regulations. Also, no studies have observed almond consumption in prediabetes adults in India.

### Strength and Limitations

To our knowledge, the present study will be the first of its kind of a field-based trial in rural settings of India to assess the effect of almond consumption versus traditional cereal pulse-based snacks on glycemic control and gut health of prediabetes adults. As such, this work has the potential to expand the current understanding of how regular consumption of almonds may help improve, reverse the prediabetes condition, and improve gut health. The compliance management is crucially designed by performing an intervention on one-to-one closed supervision, assessment of serum tocopherol levels, and daily follow up of any adverse events throughout the intervention. In India, where diabetes is known to increase disability-adjusted life years, it is crucial to establish nutrition interventions that can help prevent the disease, mainly through their impact on modifiable risk factors like hyperglycemia. The consumption of nuts aligns with this urgent need. Moreover, the upcoming metagenomic analysis and the gut metabolites like Short chain fatty acids (SCFA) and GLP 1 will significantly advance our understanding of the impact of almonds on gut health. However, a potential limitation of the study is that it will not be feasible to follow up with the participants after the intervention.

## CONCLUSION

The 16-week almond intervention randomized controlled trial could provide valuable insights into evidence-based approaches for preventing non-communicable diseases, such as diabetes, by incorporating simple lifestyle changes such as adding almonds to the diet as a powerful dietary supplement. Positive results from the study could lead to recommendations to replace commonly consumed cereal and pulse-based snacks with nuts, resulting in improved regulation of blood glucose levels and enhanced gut health.

## Data Availability

All data produced in the present work are contained in the manuscript

## Funding

This work is funded by Almond Board of California, CA, USA

## Disclaimer

This funding source has got no role in design of the study and will have no role in analysis and interpretation of the data.

## Acknowledgements

The gut microbiome and GLP-1 analysis will be performed by the Medgenome Laboratories, Bengaluru, India. The Short Chain Fatty Acid (SCFA) analysis will be performed in the Sophisticated Instrumentation Centre for Applied and Research Training (SICART) Laboratories and GSFC Research and Development Laboratories,Gujarat, India.

## Authors Contribution

RV, NC and PK were the co-applicants on the grants application and are primarily involved in study design. RV is the lead applicant and principal investigator of the study. NC, PK, SN and BP contributed to the method development and writing and development of the study protocol.

## Notes

### Competing Interest Statement

The authors have declared no competing interest.

### Clinical Trial

CTRI/2023/03/050421

## REFERENCES

1. Saeedi P, Petersohn I, Salpea P et al. IDF Diabetes Atlas Committee. Global and regional diabetes prevalence estimates for 2019 and projections for 2030 and 2045: Results from the International Diabetes Federation Diabetes Atlas, 9th edition. Diabetes Res Clin Pract. 2019;157:107843. doi: 10.1016/j.diabres.2019.107843. Epub 2019 Sep 10. PMID: 31518657.

2. Unnikrishnan R, Anjana RM, Mohan V. Diabetes in South Asians: is the phenotype different? Diabetes. 2014;63(1):53–5. doi: 10.2337/db13-1592. PMID: 24357697.

3. Mohan V, Sharp PS, Cloke HR et al. Serum immunoreactive insulin responses to a glucose load in Asian Indian and European type 2 (non-insulin-dependent) diabetic patients and control subjects. Diabetologia. 1986;29(4):235–7. doi: 10.1007/BF00454882. PMID: 3519338.

4. Mohan V, Ruchi V, Gayathri R, Bai MR, Sudha V, Anjana RM, Pradeepa R. Slowing the diabetes epidemic in the World Health Organization South-East Asia Region: the role of diet and physical activity. WHO South East Asia J Public Health. 2016;5(1):5–16. doi: 10.4103/2224-3151.206554. PMID: 28604391.

5. Gurung M, Li Z, You H et al. Role of gut microbiota in type 2 diabetes pathophysiology. EBioMedicine.2020;51:102590. doi: 10.1016/j.ebiom.2019.11.051. Epub 2020 Jan 3. PMID: 31901868; PMCID: PMC6948163.

6. Bielka W, Przezak A, Pawlik A. The Role of the Gut Microbiota in the Pathogenesis of Diabetes. Int J Mol Sci. 2022;23(1):480. doi: 10.3390/ijms23010480. PMID: 35008906; PMCID: PMC8745411.

7. Choo JM, Tran CD, Luscombe-Marsh ND et al. Almond consumption affects fecal microbiota composition, stool pH, and stool moisture in overweight and obese adults with elevated fasting blood glucose: A randomized controlled trial. Nutr Res. 2021;85:47–59. doi: 10.1016/j.nutres.2020.11.005. Epub 2020 Nov 20. PMID:33444970.

8. Ukhanova M, Wang X, Baer DJ et al. Effects of almond and pistachio consumption on gut microbiota composition in a randomised cross-over human feeding study. Br J Nutr. 2014 Jun 28;111(12):2146–52. doi: 10.1017/S0007114514000385. Epub 2014; 111, 2146–2152. PMID: 24642201.

9. Holscher HD, Taylor AM, Swanson KS et al. Almond Consumption and Processing Affects the Composition of the Gastrointestinal Microbiota of Healthy Adult Men and Women: A Randomized Controlled Trial. Nutrients. 2018;10(2):126. doi: 10.3390/nu10020126. PMID: 29373513; PMCID: PMC5852702.

10. Dhillon J, Li Z, Ortiz RM. Almond Snacking for 8 wk Increases Alpha-Diversity of the Gastrointestinal Microbiome and Decreases Bacteroides fragilis Abundance Compared with an Isocaloric Snack in College Freshmen. Curr Dev Nutr. 2019;3(8):1–9. doi: 10.1093/cdn/nzz079. PMID: 31528836; PMCID: PMC6736066.

11. Burns AM, Zitt MA, Rowe CC et al. Diet quality improves for parents and children when almonds are incorporated into their daily diet: a randomized, crossover study. Nutr Res. 2016;36(1):80–9. doi: 10.1016/j.nutres.2015.11.004. Epub 2015 Nov 10. PMID: 26773784.

12. IIPS. National Family Health Survey-5 2019–2020. Government of India. 2020.

13. Chan AW, Tetzlaff JM, Altman DG et al. SPIRIT 2013 statement: defining standard protocol items for clinical trials. Ann Intern Med. 2013;158(3):200–7. doi: 10.7326/0003-4819-158-3-201302050-00583. PMID: 23295957; PMCID: PMC5114123.

14. Mohan V, Sandeep S, Deepa M et al. A diabetes risk score helps identify metabolic syndrome and cardiovascular risk in Indians - the Chennai Urban Rural Epidemiology Study (CURES-38). Diabetes Obes Metab. 2007;9(3):337–43. doi: 10.1111/j.1463-1326.2006.00612.x. PMID: 17391160.

15. Rajput R, Garg K, Rajput M. Prediabetes Risk Evaluation Scoring System [PRESS]: A simplified scoring system for detecting undiagnosed Prediabetes. Prim Care Diabetes. 2019;13(1):11–15. doi: 10.1016/j.pcd.2018.11.011. Epub 2018 Dec 10. PMID: 30545792.

16. Kendall JM. Designing a research project: randomised controlled trials and their principles. Emerg Med J. 2003;20(2):164–8. doi: 10.1136/emj.20.2.164. PMID: 12642531; PMCID: PMC1726034.

17. Pastors JG, Warshaw H, Daly A et al. The evidence for the effectiveness of medical nutrition therapy in diabetes management. Diabetes Care. 2002;25(3):608–13. doi: 10.2337/diacare.25.3.608. PMID: 11874956.

18. GBD 2019 Diseases and Injuries Collaborators. Global burden of 369 diseases and injuries in 204 countries and territories, 1990-2019: a systematic analysis for the Global Burden of Disease Study 2019. Lancet. 2020;396(10258):1204–1222. doi: 10.1016/S0140-6736(20)30925-9. Erratum in: Lancet. 2020 Nov 14;396(10262):1562. PMID: 33069326; PMCID: PMC7567026.

19. Dreher ML. A Comprehensive Review of Almond Clinical Trials on Weight Measures, Metabolic Health Biomarkers and Outcomes, and the Gut Microbiota. Nutrients. 2021;13(6):1968. doi: 10.3390/nu13061968. PMID: 34201139; PMCID: PMC8229803.

20. Carter S, Hill AM, Yandell C et al. Study protocol for a 9-month randomised controlled trial assessing the effects of almonds versus carbohydrate-rich snack foods on weight loss and weight maintenance. BMJ Open. 2020;10(7):e036542. doi: 10.1136/bmjopen-2019-036542. PMID: 32690523; PMCID: PMC7371143.

21. Madan J, Desai S, Moitra P et al. Effect of Almond Consumption on Metabolic Risk Factors-Glucose Metabolism, Hyperinsulinemia, Selected Markers of Inflammation: A Randomized Controlled Trial in Adolescents and Young Adults. Front Nutr. 2021;8:63–68. doi: 10.3389/fnut.2021.668622. PMID: 34249987; PMCID: PMC8264510.

22. Gulati S, Misra A, Pandey RM. Effect of Almond Supplementation on Glycemia and Cardiovascular Risk Factors in Asian Indians in North India with Type 2 Diabetes Mellitus: A 24-Week Study. Metab Syndr Relat Disord. 2017;15(2):98–105. doi: 10.1089/met.2016.0066. Epub 2017 Jan 4. PMID: 28051354; PMCID: PMC5333560.

23. Gayathri R, Abirami K, Kalpana N et al. Effect of almond consumption on insulin sensitivity and serum lipids among Asian Indian adults with overweight and obesity-A randomized controlled trial. Front Nutr. 2023;9:1055923. doi: 10.3389/fnut.2022.1055923. PMID: 36704786; PMCID: PMC9873375.

